# Fetal spina bifida associates with dysregulation in nutrient-sensitive placental gene networks: findings from a matched case-control study

**DOI:** 10.1101/2023.01.25.23285010

**Authors:** Marina White, Jayden Arif-Pardy, Tim Van Mieghem, Kristin L Connor

**Affiliations:** Health Sciences, Carleton University, Ottawa, ON K1S 5B6, Canada; Department of Obstetrics and Gynaecology, Mount Sinai Hospital, Toronto, ON M5G 1X5, Canada

**Keywords:** Spina bifida, neural tube defects, placenta, transcriptome, network analysis, nutrient-gene interactions

## Abstract

To improve outcomes of fetuses with spina bifida (SB), better knowledge is needed on the molecular drivers of SB and its comorbidities. We have recently shown in historical data that SB often associates with reduced fetal growth. We here use placental transcriptome sequencing and a novel nutrient-focused analysis pipeline to determine whether this association is due to placental dysfunction. We show that fetuses with SB have dysregulation in placental gene networks that play a role in nutrient transport, branching angiogenesis, and immune/inflammatory processes. Several of these networks are sensitive to multiple micronutrients, other than the well-known folic acid, and this deserves further investigation. An improved understanding of placental phenotype in fetuses with SB may help identify novel mechanisms associated with SB and its comorbidities, and reveal new targets to improve fetal outcomes in this population.

## Introduction

Risk factors for neural tube defects (NTDs) include nutritional factors^1-4^, genetic susceptibilities, and immune and metabolic dysregulation^5-7^, yet only ∼50% of NTDs are prevented by current public health approaches^8^. This may be in part because the primary approach to reduce NTD incidence, improving maternal periconceptional folate status, only focuses on one potential mechanism. An improved understanding of other drivers of NTDs, and of how multiple risk factors may cooccur to increase the risk of NTDs, could help prevent the over 300,000 NTDs that affect newborns globally each year, and reduce morbidity risk in children with NTDs^9^.

The risk factors known to associate with NTDs are likely to also disrupt proper placental growth and development, which may in part explain why fetuses with NTDs have an increased risk of fetal growth restriction and early birth^10-15^. Optimal placentation, beginning during the first week post-fertilization and continuing across pregnancy^16^, relies on proper immune signaling and cells^17^ and bioavailable nutrients^18,19^. Thus, an early pregnancy environment permissive of NTDs may also associate with placental maldevelopment and dysfunction, which could drive comorbidities in fetuses with NTDs^10-13^. We have previously shown in a historical cohort that fetuses with NTDs have an increased risk of altered placental structure/pathology^12^. However, a more comprehensive analysis of placental function in pregnancies with fetal NTDs may reveal insights into functional contributors to comorbidities in these pregnancies, and novel gene signatures associated with NTDs.

To better understand how placental function is altered in fetuses with NTDs, we used data from a case-control study to determine how the placental transcriptome differs in fetuses with isolated spina bifida (SB; the most common NTD; cases), compared to fetuses without any congenital anomalies (controls). We hypothesised that placental transcriptome profiles in cases would be distinct from controls, especially in cases with poorer growth outcomes, and that dysregulation would be evident in gene pathways critical for fetoplacental growth. We also hypothesised that multiple nutrient-sensitive gene networks would be dysregulated in cases. By improving our understanding of the functional placental gene networks dysregulated in fetuses with NTDs, we can identify novel mechanisms associated with SB and its comorbidities, and new targets for improving fetoplacental outcomes.

## Methods

### Ethics

This study was approved by the Carleton University Research Ethics Board (108884 and 106932) and the Mount Sinai Hospital Research Ethics Board (17-0028-E and 17-0186-E).

### Population and study design

We conducted a case-control study. Pregnant people carrying a fetus with open SB (cases) were prospectively recruited from the Fetal Medicine Unit at Mount Sinai Hospital, Toronto in the early 3^rd^ trimester of pregnancy. Fetuses with structural anomalies outside the spina bifida spectrum and those with genetic anomalies were excluded. Pregnant people carrying a fetus without congenital anomalies were recruited from our low-risk pregnancy unit (SB controls) around 25 weeks’ gestation. For this placental study, placentae were collected from 12 cases and 10 SB controls. A second control group, matched to cases for gestational age at delivery, was identified to account for differences in gestational age at birth between cases and SB controls. These controls delivered preterm (PT controls; n=12) but had no other known pregnancy complications. PT controls were matched 1:1 with cases for GA at delivery and maternal pre-pregnancy body mass index (BMI) classification.

### Maternal clinical, demographic, and dietary characteristics

Maternal clinical data, medical and pregnancy history and a dietary recall questionnaire data (Automated Self-Administered 24-hour Recall™, specific to the Canadian population [ASA24-Canada-2016]^20^) were collected at recruitment for cases and SB controls.

Estimated average requirements (EARs), estimated energy requirement (EER) and tolerable upper intake levels (TULs) were calculated using the Institute of Medicine’s guidelines for pregnant people^21^. EARs represent the median daily intake amount predicated to provide nutritional adequacy for half of the healthy individuals in a specific life-stage. EER represents the average daily energy intake predicted to maintain energy homeostasis. As measures of physical activity level were not available for the participants in this study, a coefficient of 1 (equivalent to ‘sedentary’, or typical daily living activities) was applied uniformly in EER calculations^22^. TULs represent the maximum daily intake level of a nutrient that is likely to be safe for an individual.

Healthy Eating Index (HEI)-2015 scores assess how well maternal dietary intake matched recommendations from established dietary guidelines, and were calculated using the simple HEI scoring algorithm method to evaluate participant’s dietary adequacy^23^. A total HEI score was calculated to classify the diet as “good” (total score >80), “needs improvement” (total score 51-80), or “poor” (total score <51)^24^.

Dietary inflammatory index (DII) scores were used to estimate the inflammatory potential of each participant’s diet, and were calculated based on 29 food parameters (27 nutrients and 2 food items)^25^. A higher DII score corresponds to a diet with more pro-inflammatory potential.

### Infant birth outcomes

At delivery, data on pregnancy outcomes (termination of pregnancy, gestational age at delivery) and infant birthweight (transformed to z-scores using Canadian growth standards^26^) were collected.

### Placental collection and processing

#### Sample collection

Placental biopsies were collected and processed by the Research Centre for Women’s and Infants’ Health (RCWIH) immediately after delivery. Placental samples were collected from each placental quadrant (≥ 1.5 cm away from the edge and centre of the placental disk), and snap frozen in liquid nitrogen and stored at -80°C until further processing^27^.

#### RNA extraction and expression profiling

RNA was extracted from whole placental homogenates using the Qiagen RNeasy Plus Mini Kit (Qiagen, Toronto, Canada) according to the manufacturer’s protocol. RNA quality control and microarray sequencing were carried out by Genome Québec (Montreal, Canada). RNA quality and quantity were checked using the Agilent Bioanalyzer system (Agilent Bioanalyzer 2100, Santa Clara, USA; Supplementary Table S1). Placental transcriptome profiles for mRNA, lncRNA and miRNA were determined using the Clariom™ D microarray (human) and the GeneChip^®^ WT Pico Kit in two batches: SB controls and cases in September 2020, and PT controls in September 2021. Batch effect was addressed in data analysis.

### Microarray analysis

#### Gene-level transcriptome analysis

Raw expression files (.CEL files) were processed in Transcription Analysis Console software (version 4.0.2). Data were summarised using the signal space transformation algorithm and normalised using the robust multiple-array average method^28^. Differentially expressed genes (DEGs) from cases compared to PT controls, and cases compared to SB controls were identified using the eBayes method, after false discovery rate (FDR) correction (Benjamini-Hochberg method^29^) at *q* value <0.05 and an absolute log fold change (FC)≥ 2. The overlap between the two DEG lists (i.e., genes that were differentially expressed *both* in cases compared to PT controls and in cases compared to SB controls) was identified and represents the subset of genes that remained differentially expressed in cases, after accounting for possible variation due to gestational age at delivery (accounted for in the cases vs. PT control comparison) or the timing of sample collection and transcriptome sequencing (accounted for in the cases vs. SB control comparison). The list of overlapping DEGs identified was carried forward in subsequent analyses.

To understand the functional gene networks associated with case placental phenotype, we used a multi-level analysis approach to evaluate gene expression signatures at the gene, pathway, and network levels, and to identify nutrient-sensitive gene networks. First, functional analysis of DEGs was performed using Gene Ontology (GO) terms and the Protein ANalysis THrough Evolutionary Relationships (PANTHER) classification system (v16.0)^30,31^. To determine whether patterns of differential gene expression in cases differed according to placental location, DEGs with known cell-specific expression at the maternal-fetal interface were also identified (expression threshold ≥ 1; PlacentaCellEnrich) and characterized according to known expression patterns in the placenta^32,33^. Lastly, to determine the extent to which DEGs in cases are sensitive to, or interact with, nutrients, DEGs that coded for micronutrient-dependent proteins (i.e., proteins with ≥ 1 micronutrient cofactor) were identified and used to construct DEG-nutrient interaction networks^34,35^.

#### Pathway-level transcriptome analysis

To identify biological processes and pathways that may associate with placental phenotype in cases, geneset enrichment analysis (GSEA; v4.2.3) was used to identify positively- and negatively-enriched genesets in cases compared to PT and SB controls, following recommended methods^36^. In brief, GSEA was run twice: once to assess geneset enrichment in the ranked gene list resulting from the cases vs. PT controls comparison, and once to assess geneset enrichment in the ranked gene list resulting from the cases vs. SB controls comparison. The same settings were applied for both analyses: A curated human geneset file was used to define genesets pertaining to biological pathways, which excluded annotations inferred from electronic annotation^36^. Geneset size filters (minimum=15, maximum=500) resulted in the filtering out of 17739/18563 genesets. A weighted scoring scheme was used, and 1000 permutations were run^36^. As with gene-level transcriptome analysis, the overlap between positive- or negatively enriched geneset lists from the two comparisons (cases compared to PT controls and cases compared to SB controls), was identified. The resulting list, which featured genesets positively or negatively enriched in cases compared to *both* PT and SB controls, was carried forward for interpretations and visualizations.

#### Network-level transcriptome analysis

Next, we constructed regulatory networks for DEGs in cases, to identify master regulators (microRNAs [miRNAs] and transcription factors [TFs]) of genes that were dysregulated in cases. Transcripts encoding miRNAs that were differentially expressed in cases compared to both control groups were identified from the DEG list and used to construct miRNA regulatory networks for DEGs in cases. Files containing miRNA-target gene network data (miRNA targetomes), limited to experimentally validated miRNA-target interactions (miRTarBase)^37,38^, were obtained from miRWalk (v04_2022)^37^. The built-in GSEA function in miRWalk was also used to determine whether the differentially expressed miRNAs were known to interact with, or be sensitive to, nutrient-related gene pathways or biological processes^37^.

We also used iRegulon (v1.3) to identify TFs predicted to co-regulate DEGs in cases (compared to PT and SB controls)^39^. Enriched TF binding motifs, or CHIP-seq peaks (derived from ChIP-seq data sets, also referred to as tracks), were identified in the regulatory sequences in the 20 kb (kilobase; 1000 base pairs of DNA) surrounding the transcription start site (TSS) of each DEG (from −10 kb to +10 kb of the TSS; default search parameters, enrichment score ≥ 3.0)^39^. TFs with nutrient cofactors were also identified and used to construct nutrient-dependent TF-DEG regulatory networks^34^.

#### Subgroup analysis of placental transcriptome profiles in cases with poor growth

Subgroup analysis was done to explore gene expression signatures in the placenta that may associate with poor fetal growth in cases. Cases who had both a low birthweight (LBW; <2500 g), and birthweight percentile <40^th^ for GA at delivery, were classified as LBW cases (n=6). LBW is a leading cause of neonatal mortality and morbidity^40^, and a birthweight <40^th^ percentile has been associated with increased risk of still birth and suboptimal neurodevelopmental outcomes^41,42^. This cut off was chosen to identify and understand placental gene expression changes in cases who, despite not having clinical growth restriction, may still be at an increased risk of subsequent comorbidities. Five cases with normal birth weight (NBW; ≥ 2500 g) or birthweights ≥ 40^th^ percentile were classified as NBW cases. The analysis steps described above at the gene-, pathway- and regulatory network-levels, were repeated for LBW cases (n=6) vs. PT and SB controls (n=21), and NBW cases (n=5) vs. PT and SB controls (n=21), to identify placental transcriptomic signatures that were unique to cases with suboptimal growth outcomes.

#### Microarray data visualizations

DEGs were visualised in volcano plots (VolcaNoseR2^43^) and heatmaps (Heatmapper^44^, JMP [v16.0, SAS Institute Inc, Cary, NC]). Cytoscape (v3.8.2) was used to construct GSEA enrichment maps, nutrient-gene interaction networks and TF- and miRNA-DEG regulatory networks^36,45^.

### Statistical analysis

Maternal demographic, clinical and dietary data were analysed and visualised using JMP (v16.0, SAS Institute Inc, Cary, NC). Associations between study group (PT controls, SB controls and cases) and maternal data and infant birth outcomes were tested using one-way Analysis of Variance (ANOVA; for parametric data) and the Wilcoxon Rank Sum test (for non-parametric data). Differences in infant birth outcomes between study groups were also assessed in adjusted analyses, using linear models (continuous variables) and Tukey’s HSD post-hoc test (to assess between-group differences), or nominal logistic regression models (categorical variables) and Likelihood Ratio Chi Square test. Adjusted models included infant sex (male, female; categorical) and gestational age at delivery (20-41 weeks; continuous). Lastly, we evaluated associations between maternal dietary recall data and DEGs using Spearman’s rank correlation test (data are Spearman’s rank correlation coefficient [rho; ρ] with raw and false discovery rate [FDR]-adjusted p values [q-value]) and principal components analysis. Continuous data are reported as median (IQR) with p values from ANOVA/Wilcoxon (unadjusted) and multiple linear regression models (adjusted), and categorical data are n (%) with p values from Likelihood Ratio Chi Square test. Significance was set at p<0.05 (for group differences in maternal demographic, clinical and dietary data, and infant outcome data) or q<0.05 (for associations between maternal dietary recall data and DEGs).

## Results

### Clinical cohort characteristics

Baseline characteristics are documented in Table 1. As expected from the study design, all 3 groups were highly similar. Further pregnancy follow-up and delivery outcomes are presented in Table 2. Of note, 6 cases underwent *in utero* fetal spina bifida closure. Six case pregnancies (50%) and one SB control (10%) underwent pregnancy termination or delivered a stillborn fetus. PT controls were more likely than SB controls and cases to be male (Table 2). There were no differences in gestational age at delivery between cases and PT controls, however, both groups had a shorter average gestation length than SB controls.

**Table 1.**
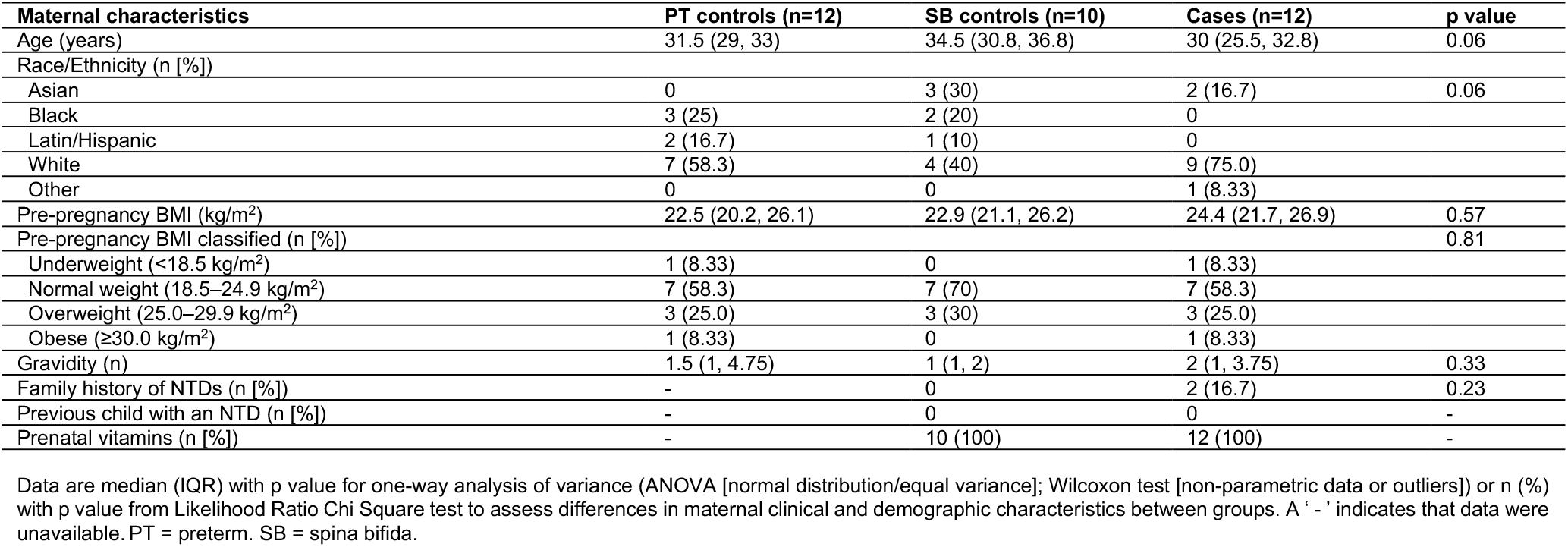
Maternal clinical and demographic characteristics.

| Maternal characteristics | PT controls (n=12) | SB controls (n=10) | Cases (n=12) | p value |
| --- | --- | --- | --- | --- |
| Age (years) | 31.5 (29, 33) | 34.5 (30.8, 36.8) | 30 (25.5, 32.8) | 0.06 |
| Race/Ethnicity (n [%]) |  |  |  |  |
| Asian | 0 | 3 (30) | 2 (16.7) | 0.06 |
| Black | 3 (25) | 2 (20) | 0 |  |
| Latin/Hispanic | 2 (16.7) | 1 (10) | 0 |  |
| White | 7 (58.3) | 4 (40) | 9 (75.0) |  |
| Other | 0 | 0 | 1 (8.33) |  |
| Pre-pregnancy BMI (kg/m<sup>2</sup>) | 22.5 (20.2, 26.1) | 22.9 (21.1, 26.2) | 24.4 (21.7, 26.9) | 0.57 |
| Pre-pregnancy BMI classified (n [%]) |  |  |  | 0.81 |
| Underweight (<18.5 kg/m<sup>2</sup>) | 1 (8.33) | 0 | 1 (8.33) |  |
| Normal weight (18.5–24.9 kg/m<sup>2</sup>) | 7 (58.3) | 7 (70) | 7 (58.3) |  |
| Overweight (25.0–29.9 kg/m<sup>2</sup>) | 3 (25.0) | 3 (30) | 3 (25.0) |  |
| Obese (≥30.0 kg/m<sup>2</sup>) | 1 (8.33) | 0 | 1 (8.33) |  |
| Gravidity (n) | 1.5 (1, 4.75) | 1 (1, 2) | 2 (1, 3.75) | 0.33 |
| Family history of NTDs (n [%]) | - | 0 | 2 (16.7) | 0.23 |
| Previous child with an NTD (n [%]) | - | 0 | 0 | - |
| Prenatal vitamins (n [%]) | - | 10 (100) | 12 (100) | - |

Data are median (IQR) with p value for one-way analysis of variance (ANOVA [normal distribution/equal variance]; Wilcoxon test [non-parametric data or outliers]) or n (%) with p value from Likelihood Ratio Chi Square test to assess differences in maternal clinical and demographic characteristics between groups. A '-' indicates that data were unavailable. PT = preterm. SB = spina bifida.

**Table 2.** Fetal spina bifida characteristics and infant outcomes at delivery.

|  | Preterm controls (n=12) | SB controls (n=10) | Cases (n=12) | Unadjusted p value | Adjusted p value <sup>a</sup> |
| --- | --- | --- | --- | --- | --- |
| **Spina bifida characteristics** |  |  |  |  |  |
| Spina bifida starting level (n [%]) |  |  |  | - | - |
| L1-2 | - | - | 1 (8.33) |  |  |
| L3-4 | - | - | 6 (50.0) |  |  |
| L5-S1 | - | - | 4 (33.3) |  |  |
| Low sacral | - | - | 1 (8.33) |  |  |
| Spina bifida-associated malformations (n [%]) <sup>b</sup> |  |  |  | - |  |
| Ventriculomegaly | - | - | 7 (58.3) |  |  |
| Talipes | - | - | 7 (58.3) |  |  |
| Arnold Chiari | - | - | 11 (91.7) |  |  |
| fMMC repair (n [%]) | - | - | 6 (50) | - | - |
| **Infant outcomes at delivery** |  |  |  |  |  |
| Stillborn/TOP (n [%]) | - | 1 (10) | 6 (50) | - | - |
| Male-to-female sex ratio | 3 <sup>a</sup> | 0.25 <sup>b</sup> | 0.5 <sup>b</sup> | 0.02 | - |
| GA at birth (weeks) <sup>c</sup> | 34.1 (31.5, 34.7) <sup>a</sup> | 39.7 (38.7, 40.1) <sup>b</sup> | 32.1 (23.8, 36) <sup>a</sup> | <.0001 | <.0001 |
| <37 weeks GA at birth (n [%]) | 12 (100) | 0 | 10 (83.3) | <.0001 | - |
| Sub-categories of preterm birth (n [%]) |  |  |  | 0.3 | - |
| Moderate/late preterm (32-36 weeks GA) | 7 (58.3) | - | 4 (33.3) |  |  |
| Very preterm (28-32 weeks GA) | 3 (25.0) | - | 2 (16.7) |  |  |
| Extremely preterm (<28 weeks GA) | 2 (16.7) | - | 4 (33.3) |  |  |
| Birthweight z score | -0.05 (-0.63, 0.45) | 0.04 (-1.3, 0.43) | -0.27 (-0.69, 0.17) | 0.6 | - |

<sup>a</sup> Adjusted for infant sex and gestational age at delivery. <sup>b</sup> There was also one case who had other fetal anomalies (lemon sign, right feet dorsiflexed, bilateral indentation of frontal bones). <sup>c</sup> Adjusted for fetal sex only. Data are median (IQR) with p value for one-way analysis of variance (Unadjusted: ANOVA [normal distribution/equal variance] or Wilcoxon test [non-parametric data or outliers]; Adjusted: Multiple standard least squares regression models) or n (%) with p value from Likelihood Ratio Chi Square test (unadjusted) and nominal Logistic regression models (adjusted) to assess differences in infant delivery outcomes between groups. Group differences from adjusted post-hoc tests (Tukey HSD) are indicated by different subscript letters. No letters = no group differences. A '-' indicates that data were unavailable. L = Lumbar; S = Sacral. fMMC = fetal myelomeningocele. TOP = termination of pregnancy. SB = spina bifida. GA = gestational age.

### Case placentae have dysregulated gene expression compared to PT and SB controls

One SB control sample failed RNA quality and quantity checks, leading to the following final sample sizes for transcriptome sequencing: PT controls (n=12), SB controls (n=9) and cases (n=12). In a comparison of global placental gene expression profiles, 1705 annotated DEGs were identified in cases compared to PT controls (downregulated in cases: n=779, upregulated in cases: n=926), and 726 were identified in cases compared to SB controls (downregulated in cases: n=388, upregulated in cases: n=338; Figure 1). The overlap between these two gene lists (i.e., genes that were differentially expressed in cases compared to both PT and SB controls) was identified and carried forward for further analyses (n=391 DEG: downregulated in cases: n=213, upregulated in cases: n=178; Supplementary Figure S1).

**Figure 1.**
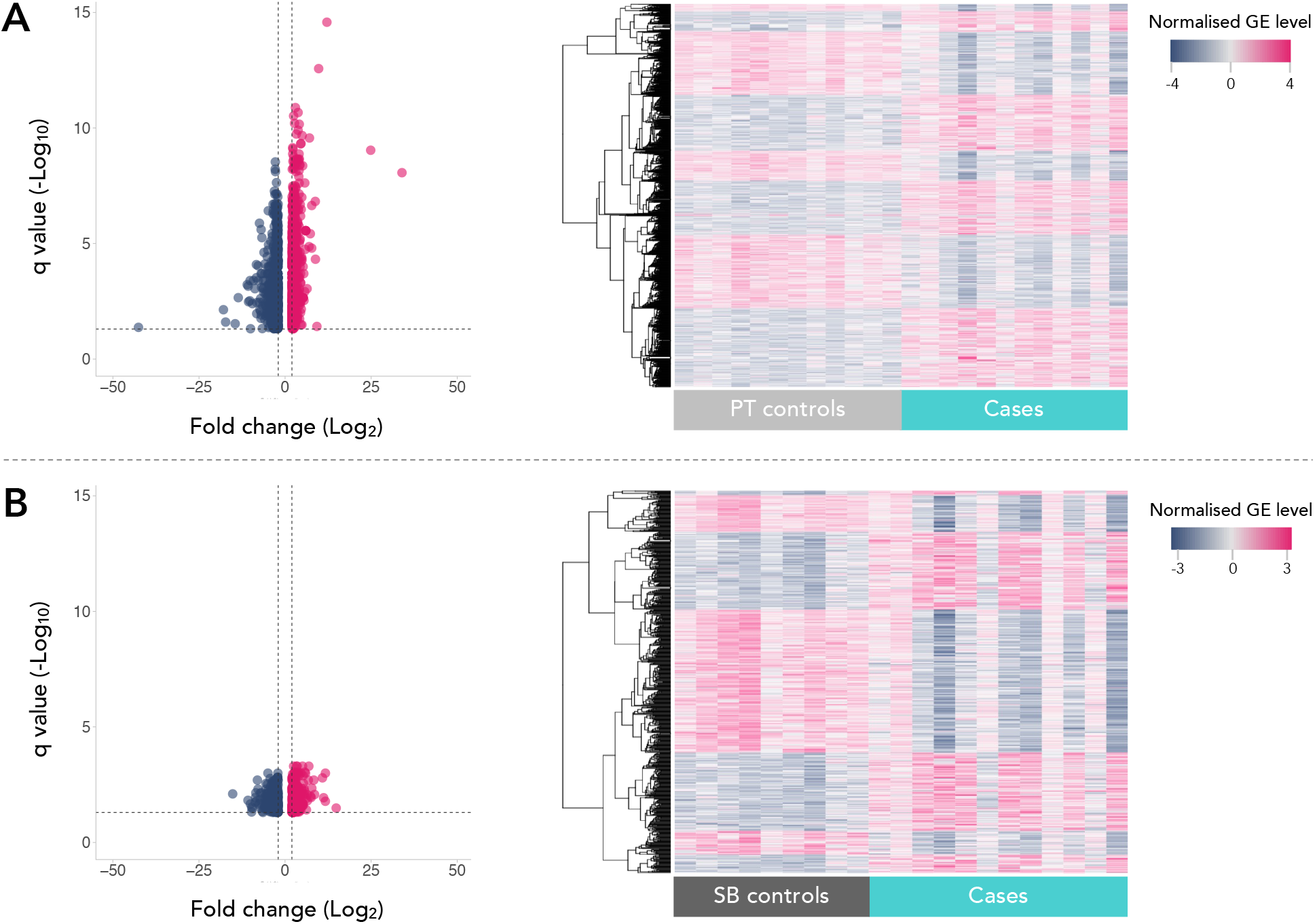
Placental gene expression differs in cases compared to (A) preterm controls and (B) spina bifida study controls. Volcano plots (left panels) and heat maps (right panels) of genes with increase and decreased expression in cases compared to (A) PT controls and (B) SB controls (clustering method: average linkage; distant measurement method: Euclidean; clustering applies to rows). Differential expression was determined at absolute fold change > 2.0 and FDR p value (q) < .05. GE = gene expression. PT controls = preterm controls. SB controls = spina bifida study controls.

The top protein classes for downregulated genes in cases were transporters, protein modifying enzymes, and defense/immunity proteins, while the top protein classes for upregulated genes were gene-specific transcriptional regulators, protein modifying enzymes and metabolite interconversion enzymes (Supplementary Figure S2). The full lists of differentially expressed genes are provided in supplementary tables S2-4.

### Case placental gene signatures vary across placental cell types

Cell-specific enrichment at the maternal-fetal interface of case placentae was identified for 110 of the 391 annotated DEGs (28.1%), of which, 69 (17.6%) had known cell-specific expression in the placenta, specifically. In general, we observed upregulation in genes with known placental cell-specific expression within the placental villi (Hofbauer cells, fetal fibroblasts), and downregulation in genes with known expression in plasma membrane cells (villous cytotrophoblast, syncytiotrophoblast, extra villous cytotrophoblast; Figure 2).

**Figure 2.**
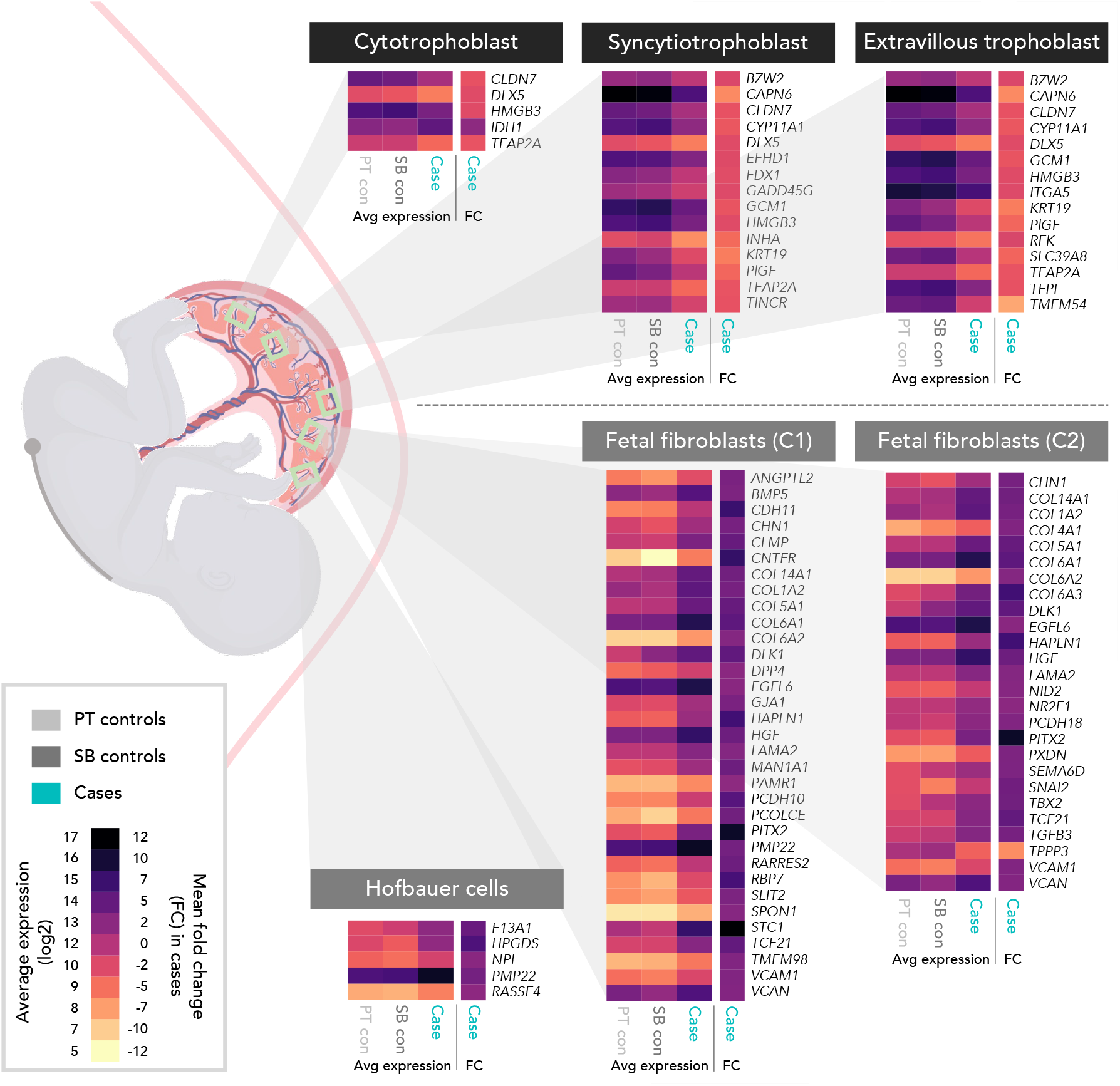
Differentially expressed genes in cases (vs. PT and SB controls) have known cell-specific enrichment in placental tissues. Heat maps show average expression for each study group (columns 1-3; left to right) and mean fold change expression in cases (vs. PT and SB controls; column 4) for genes with known cell-specific expression in trophoblast (top row of heat maps) and stromal cells (bottom row of heat maps). Placental-cell specific enrichment was evident for 69 (17.6%) DEGs in cases. Placental cell-specific expression was determined using PlacentalCellEnrich (dataset: Vento-Tormo R et al.). C1 = cluster 1. C2 = cluster 2. PT controls = preterm controls. SB controls = spina bifida study controls.

Downregulated genes in cases with known expression in placental trophoblasts included solute carrier family 39 member 8 (*SLC39A8*; transporter of essential metals), riboflavin kinase (*RFK*; enzyme involved in riboflavin [vitamin B2] utilisation), and Ferredoxin 1 (*FDX1*; involved in vitamin D metabolism). Placental growth factor (*PlGF*), a pro-angiogenic regulator of placental vasculogenesis and non-branching angiogenesis, was also downregulated in cases and has known trophoblastic expression.

Upregulated genes in cases with known expression in Hofbauer cells included hematopoietic Prostaglandin D Synthase (*HPGDS*; involved in inflammatory response regulation), while genes with known expression in fibroblasts included extracellular matrix structural proteins, hyaluronan and proteoglycan link protein 1 [*HAPLN1*], versican [*VCAN*]) and growth factors.

### Case placentae have positively and negatively enriched gene pathways

GSEA revealed 437 enriched gene pathways in cases compared to PT controls (downregulated in cases: n=124, upregulated in cases: n=313), and 528 enriched gene pathways in cases compared to SB controls (downregulated in cases: n=306, upregulated in cases: n=222; Supplementary Tables S5 and S6). There were 122 enriched gene pathways in cases compared both PT and SB controls (downregulated in cases: n=30, upregulated in cases: n=92), as shown in Figure 3A and Supplementary Figure S3.

**Figure 3.**
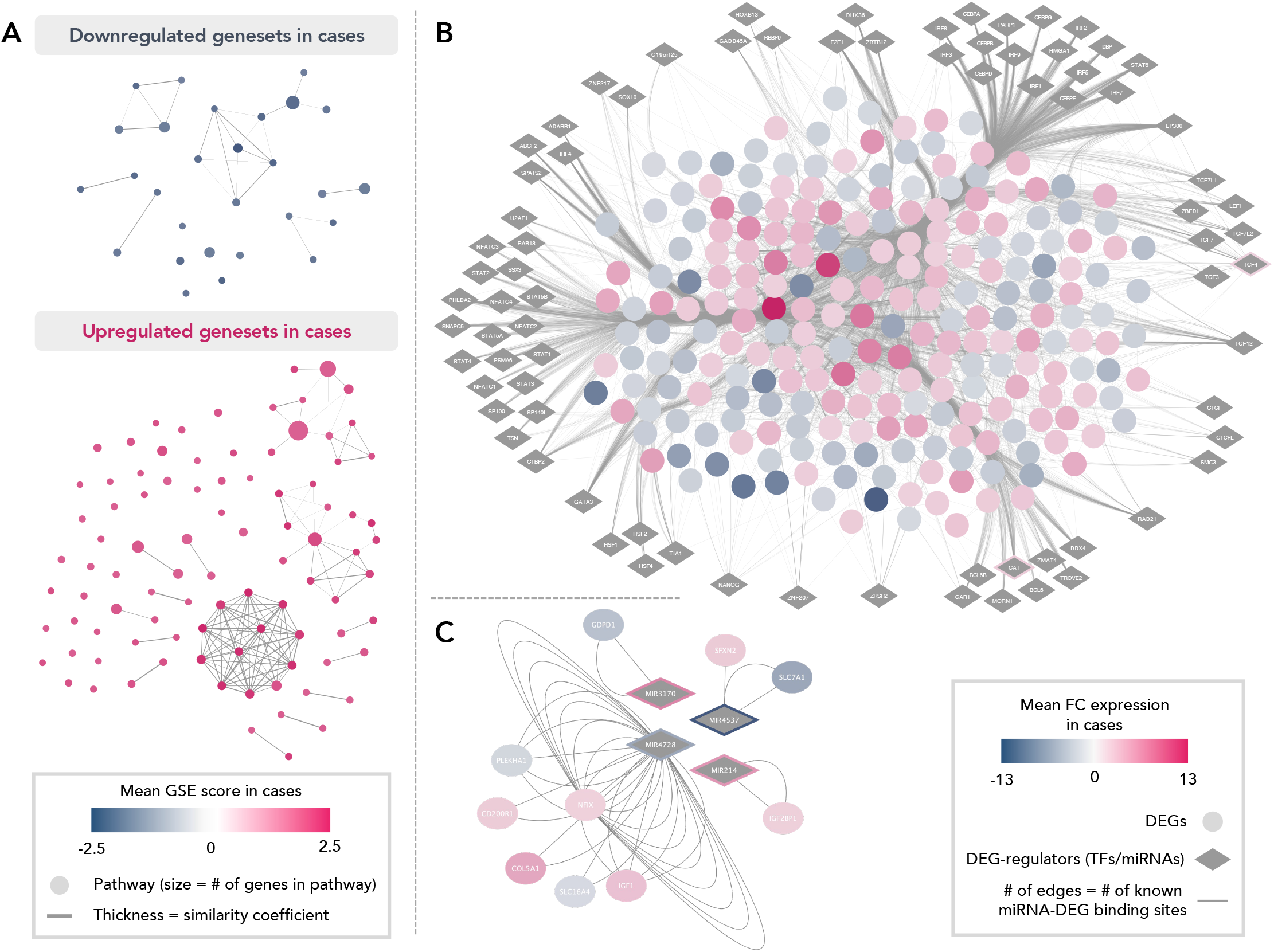
Differentially expressed genesets and transcription factors and miRNAs co-regulating differentially expressed genes in case placentae. (A) Geneset enrichment analysis revealed 122 enriched gene pathways (nodes) in cases compared to PT and SB controls (downregulated in cases: n=30 [blue nodes], upregulated in cases: n=92 [pink nodes]; for an annotated version of this figure, see Supplementary Figure S3). (B) 78 TFs (diamonds) putatively regulating 209 (53.3%) DEGs in cases. TFs are loosely grouped according to module (i.e., sets of TFs targeting the same or a similar subset of DEG). (C) Four differentially expressed miRNAs in cases (diamonds) are known to target 10 DEGs (2.6%) in cases. GSE = geneset enrichment. TF = Transcription factors. DEGs = differentially expressed genes. FC = fold change.

Downregulated genesets in cases included gene pathways related to coat protein complex II (COPII)-mediated vesicle transport from the endoplasmic reticulum (ER) to the Golgi apparatus, ER organization and signaling, protein localization (within the ER and to the cell membrane), and glycosylation. Gene pathways involved in transmembrane influx and efflux transport, including solute carrier (SLC) transport (amino acid transport) and ATP-binding cassette (ABC) transport, and lipid metabolism, were also downregulated.

Upregulated genesets included gene pathways related to immune and inflammatory processes (Interleukin 6-mediated signaling events, positive regulation of chemokine production and activated T cell proliferation, and the inflammatory response pathway) and spinal cord injury, and the largest, and most interconnected cluster of upregulated pathways in cases was involved in translation and protein synthesis. Cases also had upregulation in O-linked glycosylation-related pathways, including O-glycosylation of TSR domain-containing proteins, and defective beta-1,3-glucosyltransferase (*B3GALTL*) causes Peters plus syndrome (PpS; associates with delayed growth and neurodevelopment, cleft lip/palate^46^). Gene pathways involved in branching angiogenesis and epithelial tube formation, extracellular matrix formation (namely collagen formation), organization and signaling, as well as the Hedgehog and Wnt signaling pathways (critical regulators of cell proliferation, differentiation, and development) were also upregulated in cases.

### Differentially expressed genes in case placentae have common transcriptional regulators

At the gene regulatory network level, we identified 78 TFs putatively regulating 209 (53.3%) DEGs in cases (Figure 3B, Supplementary Table S8). The top candidate TFs (i.e., those associated with the most enriched DNA motifs/tracks in the DEG list) were TROVE domain family, member 2 (*TROVE2*; regulating 14.1% of DEGs [n=55] in cases), heat shock transcription factors (*HSF1, HSF2, HSF4*; regulating 6.7% of DEGs [n=26] in cases) zinc finger protein 207 (*ZNF207*; regulating 5.4% of DEGs [n=21] in cases), DEAD-Box helicase 4 (DDX4; regulating 19.9% of DEGs [n=78] in cases), and high mobility group AT-hook 1 (HMGA1; regulating 11.5% of DEGs [n=45] in cases). Pleckstrin Homology Like Domain Family A Member 2 (*PHLDA2*) was predicted to target the largest proportion of DEGs in cases (n=124 [31.7%]).

Two of the TFs predicted to regulate DEGs were upregulated in cases (transcription factor 4 [*TCF4*]: mean FC=2.1, mean q=0.01; and catalase [*CAT*]: mean FC=2.4, mean q=0.01). *TCF4* and *CAT* were predicted to regulate 6.7% (n=26) and 20.4% (n=80) of DEGs in cases, respectively.

We also identified nine miRNAs that were differentially expressed in cases compared to PT and SB controls (Supplementary Table S4). Collectively, these differentially expressed miRNAs targeted 10 DEGs (2.6%) in cases, however, only two of these DEG targets were among the top 50 dysregulated genes in cases (solute carrier family 7, member 1 [*SLC7A1*]: mean FC in cases = -5.8, q=0.02; collagen, type V, alpha 1 [*COL5A1*]: mean FC in cases = 4.6, q=0.003).

### Nutrient-sensitive genes and gene networks are dysregulated in case placentae

To determine the extent to which DEGs in cases were nutrient-dependent, and with which nutrients DEGs interact, we identified 42 DEGs (10.7%) in cases that coded for a protein with a nutrient cofactor (downregulated in cases: n=19, upregulated in cases: n=23). Nutrient cofactors included: calcium (for n=12 genes), B vitamins (n=7), iron/heme (n=6), metal/metal cation (n=4) and zinc (n=11; Figure 4A).

**Figure 4.**
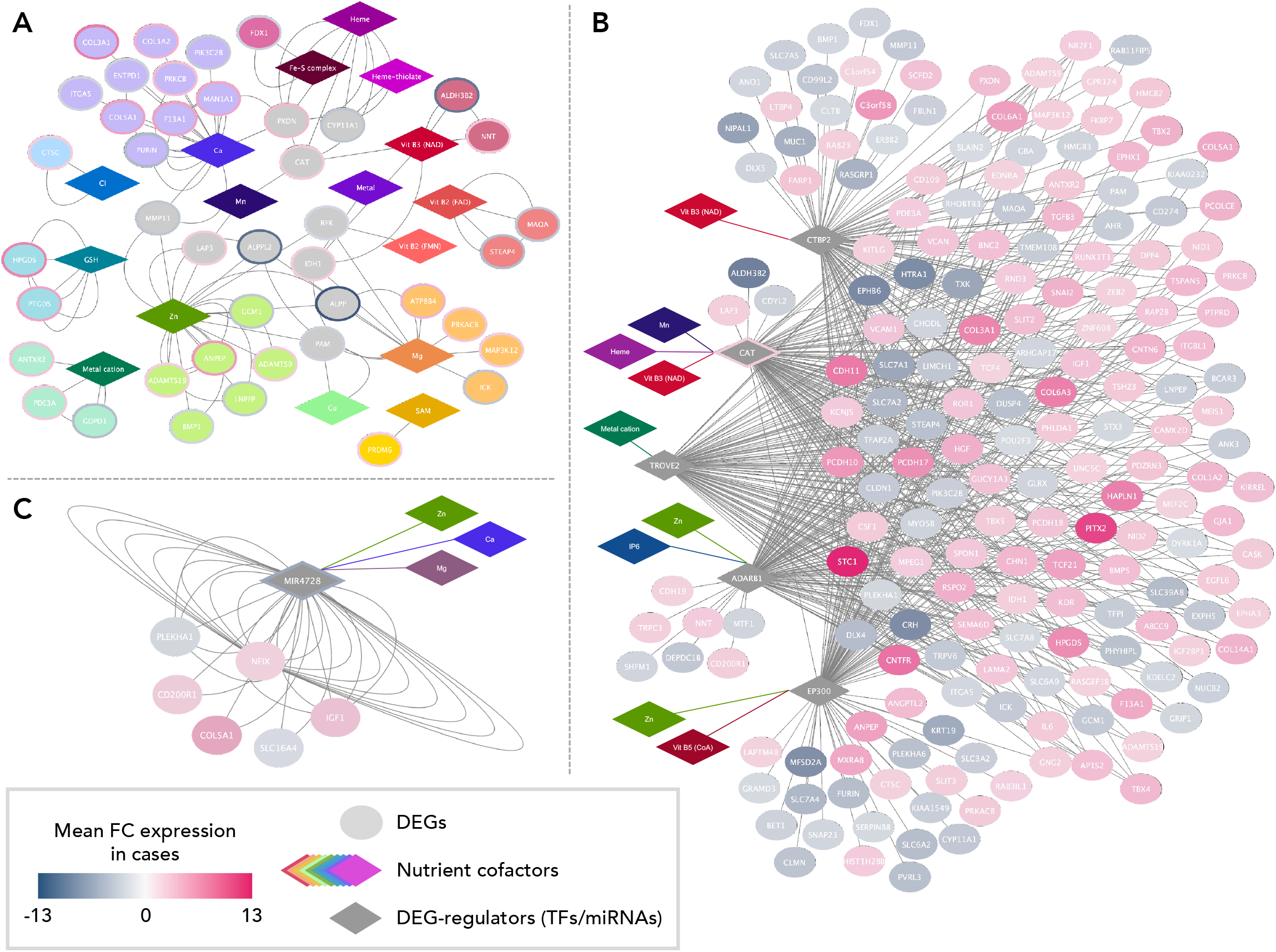
Differentially expressed genes in cases compared to PT and SB controls have nutrient cofactors (A) and are co-regulated by nutrient-sensitive TFs (B) and differentially expressed miRNAs (C). (A) DEGs in cases (circular nodes) that code for nutrient (cofactor; diamonds) dependent proteins. DEGs are coloured and clustered by their nutrient cofactors. DEGs with >1 nutrient cofactor are coloured grey. Data on nutrient-gene (protein) interactions was obtained from Scott-Boyer et al. 2016. (B) Nutrient-dependent TFs predicted to regulate DNA motifs or tracks enriched in DEG. TFs are linked to their nutrient cofactors (coloured diamonds) in the network. (C) One differentially expressed miRNA in cases is known to target DEG, and is sensitive to zinc, calcium, and magnesium activity. In panels B and C, DEGs = circle nodes, with increased (pink) or decreased (blue) expression in cases compared to PT and SB controls. DEGs = differentially expressed genes. TF = transcription factor. FC = fold change.

Next, we identified nutrient-sensitive master regulators (TFs and miRNAs) that target DEGs in cases, to determine which nutrients the dysregulated gene networks in cases were sensitive to. Of the TFs predicted to target DEG, five (6.4%) had nutrient cofactors (Figure 4B). These included C-terminal binding protein 2 (*CTBP2*; vitamin B3-dependent and a known regulator of epithelial gene expression and apoptosis^47^), *CAT* (vitamin B3-, manganese-, and heme-dependent and a key placental antioxidant^48^), *TROVE2* (metal cation-dependent and a regulator of immune and inflammatory processes^49,50^), adenosine deaminase RNA specific B1 (*ADARB1*; IP-6- and zinc-dependent with known roles in antiviral defense and immunity^50^), and E1A binding protein P300 (*EP300*; zinc- and vitamin B5-dependent with known roles in the regulation of cell cycle, transcription, and cell growth and differentiation^50^). Collectively, these nutrient-dependent TFs were predicted to target 45.9% of DEGs in cases (heme-dependent: 20.4% of DEG, manganese: 20.4% of DEG, metal cation: 14.0% of DEG, IP-6: 17.6% of DEG, vitamin B3: 35.2% of DEG, vitamin B5: 23.2% of DEG; zinc: 31.9% of DEG). One of the downregulated miRNAs in cases (*MIR4728*) was known to be sensitive to zinc-, calcium- and magnesium-activity (Figure 4C).

### Cases with poor growth and low birth weight have unique placental transcriptome profiles

We next aimed to explore associations between placental gene expression with poor fetal growth in cases. One case, whose gestational age at delivery was too young to determine birthweight percentile, was excluded from subgroup analysis, leaving 11 cases.

Global placental gene expression profiles differed more in LBW cases vs. all PT and SB controls (n=213 annotated DEGs; downregulated in LBW cases: n=148, upregulated in LBW cases: n=65; Supplementary Table S9), than NBW cases vs. all PT and SB controls (n=18 annotated DEGs, all upregulated in NBW cases; Supplementary Table S10). To characterise gene signatures unique to LBW cases, we focused on differentially expressed genes and pathways that were different in LBW, but *not* NBW, case placentae compared to PT and SB controls.

The top protein classes for genes downregulated in LBW cases only (Figure 5A; Supplementary Table S11), compared to PT and SB controls, were protein modifying enzymes and transporters, while the top protein classes for genes upregulated in LBW cases were metabolite interconversion enzymes, protein modifying enzymes, and gene-specific transcriptional regulators (Supplementary Table S12).

**Figure 5.**
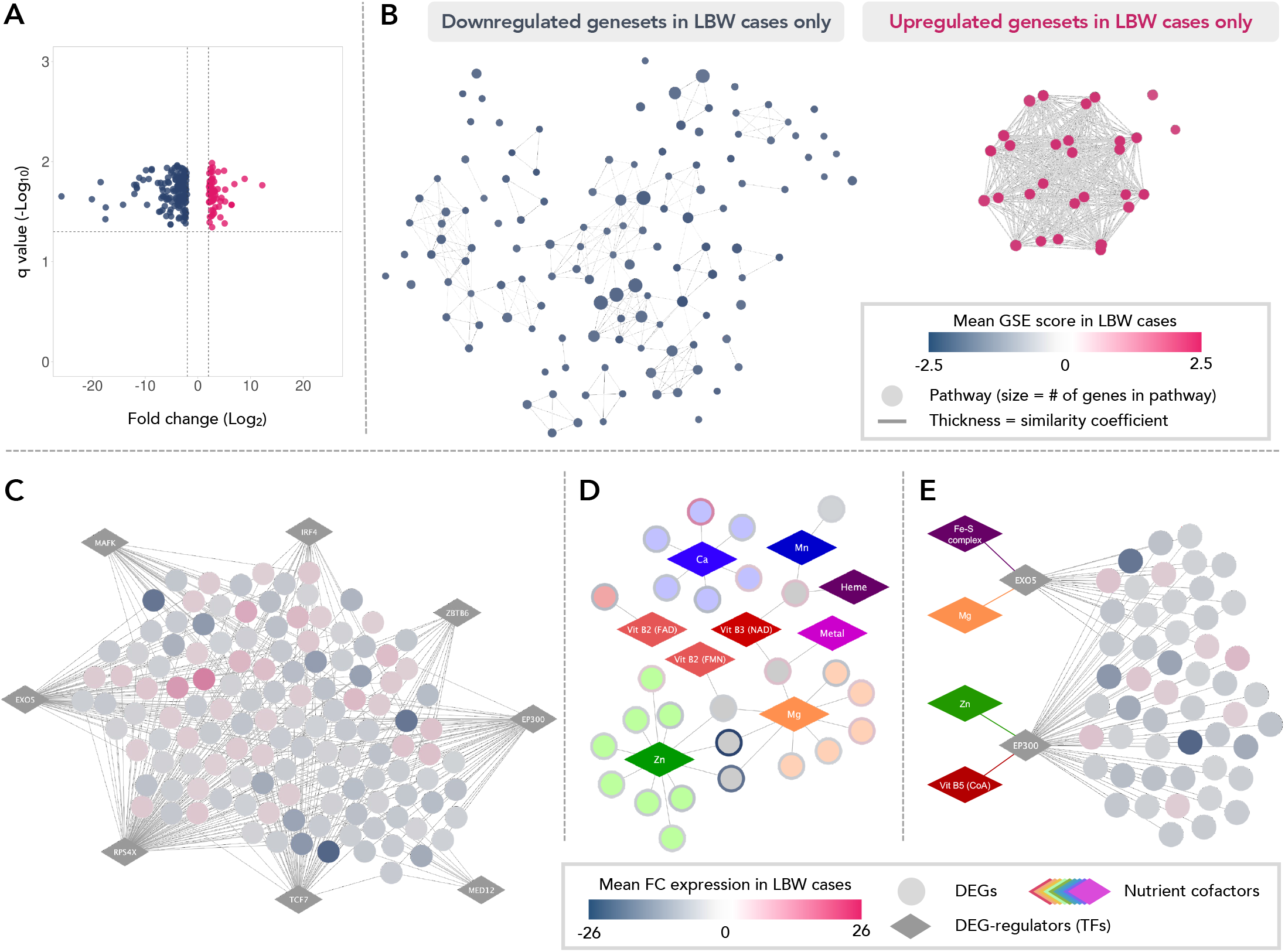
Differentially expressed placental genes, genesets and gene networks unique to cases with suboptimal growth. (A) Volcano plot of genes with increased (n=60) and decreased (n=148) expression in LBW cases only compared to PT controls and SB controls. Differential expression was determined at absolute FC>2.0 and FDR p value (q)<0.05. (B) Geneset enrichment analysis revealed 184 enriched gene pathways (nodes) in LBW cases compared to PT and SB controls (downregulated in LBW cases: n=140 [blue nodes], upregulated in LBW cases: n=44 [pink nodes]). (C) 42 TFs putatively regulating enriched DNA motifs associated with 131 (63%) DEGs unique to LBW cases were identified. Top enriched TFs (diamonds; normalised enrichment score >3) are visualised. (D) 26 (12.5%) of DEGs in LBW cases (circular nodes) code for proteins that are nutrient dependent (cofactors; diamonds). DEGs are coloured and clustered by their nutrient cofactors. DEGs with >1 nutrient cofactor are coloured grey. (E) Nutrient-dependent enriched TFs predicted to regulate DNA motifs or tracks enriched in DEGs in LBW cases. TFs are linked to their nutrient cofactors (coloured diamonds) in the network. Edges (lines) connecting nodes (circles/diamonds in B-E) indicate geneset interactions (B), TF-regulatory target links (C, E), or nutrient-gene interactions (D). LBW = low birth weight. GSE = geneset enrichment. TF = Transcription factors. DEG = differentially expressed genes. FC = fold change. PT controls = preterm controls. SB controls = spina bifida study controls.

At the pathway level, LBW case placentae had dysregulation in 154 unique genesets compared to PT and SB controls (downregulated: n=125, upregulated: n=29; Figure 5B). Consistent with GSEA results from analysis of the entire cohort of cases, LBW cases had downregulation in genesets involved in COPII-mediated vesicle transport from the ER to the Golgi apparatus, ER organization and signaling, protein localization (within the ER and to the cell membrane), and glycosylation (Figure 5B, Supplementary Table S13). LBW cases also had downregulated amino acid transport, transmembrane (ABC) transport, and lipid metabolism (Figure 5B, Supplementary Table S13). Conversely, while NBW cases also had downregulation in pathways related to COPII-mediated vesicle transport, protein localization, and amino acid transport, they were fewer in number than the changes seen in these pathways in LBW cases (15 pathways total in NBW cases vs. 56 pathways in LBW cases; Supplementary Tables S13 and 14). NBW cases did not have downregulation in lipid metabolism and transmembrane (ABC) transport, as seen in LBW cases. LBW cases had upregulation in genesets related to protein synthesis (n=27 pathways) and one apoptosis-related pathway (Figure 5B, Supplementary Tables S13 and 14).

At the gene regulatory network level, we identified 42 TFs putatively regulating enriched DNA motifs that associated with 131 (63%) DEGs unique to LBW cases (Supplementary Table S15). Seven TFs were significantly enriched, predicted to collectively regulate 127 (61.1%) DEGs unique to LBW cases (Figure 5C). The most enriched candidate TFs were interferon regulatory factor 4 (*IRF4*; a regulatory TF in human immune cell development; regulating 10.6% of DEGs [n=22] unique to LBW cases), ribosomal protein S4 X-linked (*RPS4X*; encodes a component of the ribosomal 20S subunit; regulating 48.6% of DEGs [n=101] unique to LBW cases), and transcription factor 7 (*TCF7*; a regulatory TF in innate immune system development; regulating 26.4% of DEGs [n=55] unique to LBW cases; Supplementary Table S15).

While there were four miRNAs that were differentially expressed in LBW cases, and not NBW cases, compared to PT and SB controls (Supplementary Table S11), none were known to target DEGs in LBW cases.

To determine the extent that nutrient-gene interactions may explain dysregulated gene signatures in LBW cases, we next identified 26 DEGs (12.5%) unique to LBW cases that coded for a protein with a nutrient cofactor (Figure 5D). Notably, the B vitamin-interacting genes that were dysregulated in LBW cases included genes involved in the metabolism of B vitamins (downregulated: riboflavin kinase [*RFK*; vitamin B2]^51^, alkaline phosphatase, placental [*ALPP*] and placental like 2 [*ALPPL2*; vitamin B6]^52,53^); upregulated: thiamin pyrophosphokinase 1 [*TPK1*; vitamin B1 metabolism]^51^).

Last, we identified two enriched TFs that had nutrient cofactors and targeted DEGs unique to LBW cases (Figure 5E). These TFs were exonuclease 5 (*EXO5*; iron- and magnesium-dependent with a known role in DNA repair) and E1A Binding Protein P300 (*EP300*; vitamin B5- and zinc-dependent with known roles in a transcription factor with known roles in cell growth, differentiation, and NTDs). Collectively, these two nutrient-dependent TFs were predicted to target 24.5% of DEGs unique to LBW cases (Figure 5E).

### Maternal nutrient intakes differ in case pregnancies compared to controls

In 24-hour maternal dietary recalls, mothers of cases (compared to SB controls) reported lower absolute daily intakes of choline, potassium, and vitamins A, B6, B12, C and D (Supplementary Table S16). Case mothers also met a lower overall proportion of EARs compared to SB controls (55.3% [50, 67.1] vs. 81.6% [67.1, 84.2] EARs met, p=0.01), and were more likely to fall into a lower quartile for the overall % EARs met (Figure 6A).

**Figure 6.**
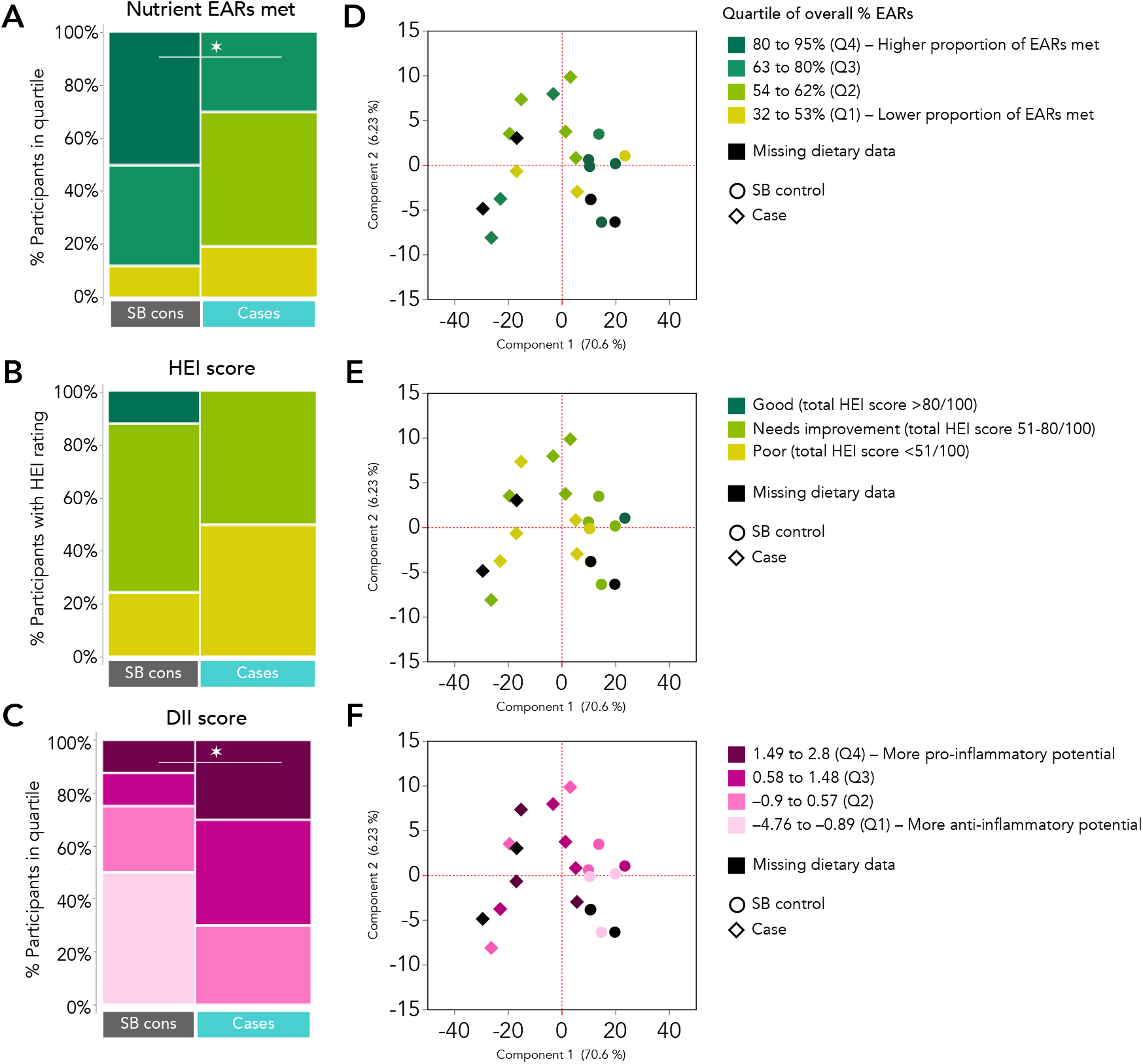
Maternal dietary intakes and differential placental gene expression patterns in cases and SB controls. (A) Mothers of cases met a lower proportion of nutrient EARs (i.e., fell in a lower quartile for % EARs met) than those of SB controls (p=0.006). (B) There were no differences between cases and SB controls for overall HEI score rating. (C) Mothers of cases had higher DII scores (i.e., fell in a higher quartile for DII score) than those of SB controls (p=0.04). (D-F) Principal component analysis of cases (diamonds) and SB controls (circles) based on the 391 differentially expressed genes in cases vs. PT and SB controls, with data representing the (D) quartile of overall % EARs met, (E) HEI score rating, and (F) DII score quartile overlayed in colouring to visualize relationships between dietary recall data and differential gene expression patterns. Width of mosaic plots (A-C) = proportional to group size. Q = quartile. EARs = estimated average requirements. HEI = health eating index. DII = dietary inflammatory index.

Mothers of cases also had lower scores for adequacy of total fruit and vegetable intakes, and intake of refined grains in moderation compared to SB controls, however, there were no group differences for total maternal HEI scores (Figure 6B and Supplementary Table S17). Lastly, compared to SB controls, mothers of cases had higher overall DII scores (0.8 [0.4, 2] vs. -1 [-2.3, 0.3], p=0.01, Supplementary Table S17), and were more likely to have a DII score in a higher quartile (Figure 6C and Supplementary Table S17).

While there were many associations between differentially expressed genes and maternal dietary intake variables, none persisted after FDR-correction (overall or within case and SB control groups; Supplementary Table S18). While there was loose clustering of cases whose mothers met 54-62% of their overall proportion of nutrient EARs, and among controls with the lowest quartile of maternal DII score, no clear separation between study participants was evident based on key dietary recall data and differential gene expression.

## Discussion

Using high throughput gene expression profiling, we identified multiple novel genes and gene regulatory networks, including those that are nutrient-sensitive, that were dysregulated in placentae of fetuses with isolated SB. Our work uncovered four major themes related to placental dysfunction in fetuses with isolated SB, advancing our understanding of SB phenotypes and placental mechanisms that may contribute to comorbidities in affected fetuses. First, we found dysregulation in gene networks that have known links to NTDs, but which have not been previously profiled in the placenta. Second, we identified new signatures in fundamental placental processes, including placental transport and growth, that were dysregulated in cases. Next, we found that several of the dysregulated placental gene networks in cases were sensitive to multiple nutrients, and that mothers of cases reported dietary intakes with lower nutritional value and higher inflammatory potential. These findings support a need to expand beyond a folic acid-centric view to understand NTD pathogenesis, phenotypes, and comorbidities. Last, we found dysregulation in gene networks related to inflammation and immune regulation in cases. Collectively, our findings suggest that the placental transcriptome is altered in fetuses with SB, and that placental dysfunction in fetuses with SB may, in part, underlie poor fetal growth, and contribute to subsequent comorbidities^54^ and increased risk of mortality^55^.

We have shown for the first time that several genes and gene pathways with known links to NTDs are dysregulated in placentae from fetuses with isolated SB. The COPII pathway, which was downregulated in case placentae, is a critical regulator of forward trafficking of proteins and lipids from the endoplasmic reticulum (ER)^56^. Disruptions in COPII-mediated vesicular ER-to-Golgi transport have been linked to SB via the role of COPII in transporting key components of the planar cell polarity (non-canonical Wnt) pathway^57-59^. Wnt signalling, as well as Hedgehog signalling, have been associated with NTDs through their regulatory role in neural tube closure^60,61^, and were both upregulated in case placentae. Notably, cases also had downregulation in genesets involved in glycosylation (a protein or lipid modification that largely occurs in the Golgi^62^), and upregulation in protein synthesis (which occurs mainly on the ribosomes of the ER), which could both be compensatory for reductions in COPII-mediated ER-to-Golgi protein transport. Lastly, *CAT* was upregulated in case placentae and identified as a putative transcriptional regulator of 20% of DEGs. *CAT* responds to oxidative stress within the cell, including ER stress that can arise due to protein accumulation^63^, and maternal CAT functional capacity may be altered in pregnancies with fetal NTDs^64^. Collectively, these findings suggest that gene expression signatures associated with NTDs may be identifiable in placentae from affected pregnancies, and that studying placental phenotype in pregnancies with fetal NTDs could provide novel insights into NTD phenotype and/or pathophysiology.

Cases also had dysregulation in placental gene networks related to placental growth, which could contribute to placental dysfunction and common SB-associated comorbidities, such as poor fetal growth. Specifically, cases had upregulation in genesets related to placental branching angiogenesis, and genes known to be expressed by Hofbauer cells and fibroblasts, which both play critical roles in placental angiogenesis^65,66^. Two insulin like growth factor (*IGF*) family genes (*IGF1* and IGF 2 mRNA binding protein 1 [*IGF2BP1*]), which have been associated with accelerated placental villous maturity in idiopathic spontaneous preterm birth^67^, were also upregulated in cases. Conversely, cases had decreased expression of two pro-angiogenic genes: *PGF* (involved in non-branching angiogenesis^68^) and Erb-B2 Receptor Tyrosine Kinase 2 (*ERBB2*; involved in trophoblast migration and differentiation)^69,70^. Collectively, these signaling alterations may have led to premature villous differentiation towards terminal villi^71,72^, which can diminish the placenta’s capacity to grow sufficiently and lead to placental dysfunction^73-75^. These transcriptomic signatures may also suggest a placental phenotype in cases that is consistent with findings in our previous work with a historical cohort, where isolated fetal NTDs associate with increased placental Hofbauer cell levels and increased risk of placental villous hypermaturity^12^. Altogether, our findings indicate that cases have altered activity in placental angiogenic genes and gene pathways, which may place them at an increased risk of placental villous maldevelopment and subsequent fetal comorbidities.

Cases also had general downregulation of genes known to be enriched in villous trophoblast cells (cytotrophoblast, syncytiotrophoblast, and extravillous trophoblast), as well as genes and genesets involved in transmembrane transport. These included ABC transport pathways, which are essential for placental efflux (defense) and the transport of nutrients (including folates, amino acids, and glucose) and inflammatory molecules, as well as SLC transport genes and pathways, which influx a wide range of nutritional and exogenous compounds^76^. More of these pathways were downregulated in cases with LBW compared to controls, rather than cases with NBW, suggesting that a potential compromise in placental barrier and nutrient transport function may underly poor growth in fetuses with SB, increasing risk of subsequent morbidities. Future studies of larger cohorts should robustly explore placental gene expression in fetuses with SB and suboptimal growth.

While maternal folate status has been the primary nutrient target for NTD prevention to-date, we found that case placentae had dysregulation in gene networks sensitive to nutrients beyond folate, including those interacting with other B vitamins, zinc, calcium, inositol, and metals/metal cations. As folate deficiency is rare in the folic acid-fortified Canadian population^77^, NTD risk may be, in part, attributable to low bioavailability of other nutritional factors; namely, one carbon metabolism cofactors (vitamins B6, B12, and choline)^2,78^. In further support of this theory, mothers of cases reported lower absolute intakes in vitamins B6, B12, and choline, but not folate, as well as vitamins C and D, and overall, met a lower proportion of their estimated nutrient requirements. Further, nearly half of DEGs in cases were putatively co-regulated by TFs dependent on nutrients with known links to NTDs, including and zinc^3,4^ and inositol^79^, and four of the five nutrient-dependent TFs (*CAT, CTBPR2, TROVE2*, and *EP300*) predicted to regulate DEGs in cases were also identified as top TFs targeting DEGs in fetuses with NTDs in our previous study^35^. Nutrient cofactors for these TFs included zinc, vitamins B3 and B5, inositol hexaphosphate, and heme. As the placenta is a rapidly developing and metabolically active organ, with considerable nutritional requirements itself throughout gestation, it is possible that low bioavailability of these nutrients may be not only permissive of fetal NTDs, but also placental maldevelopment and dysfunction in nutrient-sensitive gene networks, increasing risk subsequent offspring morbidities.

We have also shown for the first time that placentae from fetuses with isolated SB have increased expression in immune and inflammatory processes, which could increase risk of poor fetal outcomes^80^. Specifically, cases had upregulation in immune and inflammatory process gene pathways, and in genes with known expression in Hofbauer cells (including *HPGDS*, which plays an important role in inflammation response regulation^81^). As well, *ZNF207*, which was predicted to target one of the most enriched DNA modules in DEGs in cases overall, and *IRF4* and *TCF7*, two enriched TFs targeting DEGs in LBW cases specifically, all have known roles in transcriptional regulation of immune development and response^51,82,83^. Maternal immune dysregulation and increased inflammatory load are known to associate with increased risk of fetal NTDs^6,84^, and here, mothers of cases also reported dietary intakes with higher dietary inflammatory potential than SB controls. Further, following failed closure of the neural tube, ‘secondary lesion cascades’, activated in response to spinal cord injury, may induce ongoing pro-inflammatory molecular and cellular signaling alterations^85^. In this study, case placentae also had increased expression in genesets responding to spinal cord injury. Taken together, our findings suggest that placental inflammatory processes may be elevated in case pregnancies, with possible contributions from NTD-related biological processes and/or maternal dietary inflammatory potential. As placental inflammation is known to increase risk of fetal morbidity and mortality^80^, understanding whether inflammatory load is heightened in placentae from fetuses with NTDs should be a focus of further study.

Our ability to draw conclusions about the dietary recall data presented in this study are limited by the fact that only a single recall was collected, that it is known participants often underreport dietary intakes, and an objective assessment of nutritional biomarkers to validate recall results was not available^86^. Half of the cases were also delivered following pregnancy termination, whereas all controls but one were liveborn. However, as the case pregnancies that underwent termination did so at much earlier in gestation than the cases that were liveborn, it is difficult to delineate the potential effects of the termination procedure versus gestational age on placental gene expression. This is a limitation of our study, and a larger cohort would be needed to evaluate the effects of termination or liveborn with fetoscopic repair on placental phenotype in fetuses with SB. Further, while the inclusion of an additional control group (PT controls) that was gestational age-matched to cases is a strength of this study, placental dysfunction is associated with PT birth, and it is possible that the PT controls themselves may have had placental maldevelopment^15^, which could have confounded our comparisons. However, in an aim to address this limitation, we took a conservative approach when identifying the DEG list, and only considered the overlap between the two comparison lists: cases compared to SB controls (who were born full term) and cases compared to PT controls.

This is the first study to assess global placental gene expression in fetuses with isolated SB, and among the first to interrogate placental phenotype in pregnancies with fetal NTDs more broadly. Our findings increase our understanding of placental function in pregnancies with isolated fetal SB, and report multiple novel gene regulatory networks, including those that are sensitive to nutrient activity and levels, that may associate with SB phenotype and comorbidities. Our findings of dysregulated gene networks known to associate with SB pathogenesis suggests, for the first time, that functional perturbations contributing to SB may also present in placentae from these pregnancies, and potentially contribute to placental maldevelopment and dysfunction as well. Studying placental maldevelopment and dysfunction, including nutrient-gene interactions, in fetuses with isolated SB may help identify novel mechanisms associated with SB and its comorbidities, and new targets for SB prevention and improving fetal outcomes.

## Data Availability

All data produced in the present study are available upon reasonable request to the authors.

## Acknowledgments

We would like to thank the patients and families who took the time to participate in this project and generously donated their samples. We would also like to thank the staff at the Research Centre for Women’s and Infants’ Health (RCWIH) BioBank for collecting and processing the placental samples, and the staff at Genome Québec for performing the microarray sequencing. Thank you also to Vagisha Pruthi for overseeing the study recruitment, data entry and sample collection, and to Dr. Bénédicte Fontaine-Bisson for assisting with the dietary recall data analysis.

## Author contributions

Conceptualization: MW, KLC, TVM; Methodology: MW, KLC; Patient recruitment and sample collection: TVM; Formal analysis: MW, JAP; Investigation: MW, JAP, KLC; Writing – original draft preparation: MW, KLC; Writing – review and editing: MW, KLC, TVM, JAP; Supervision: KLC; Visualization: MW, KLC; Funding acquisition: KLC, TVM.

