## Supplementary Figures S1-3 for "Fetal spina bifida associates with dysregulation in nutrient-sensitive placental gene networks: findings from a matched case-control study"

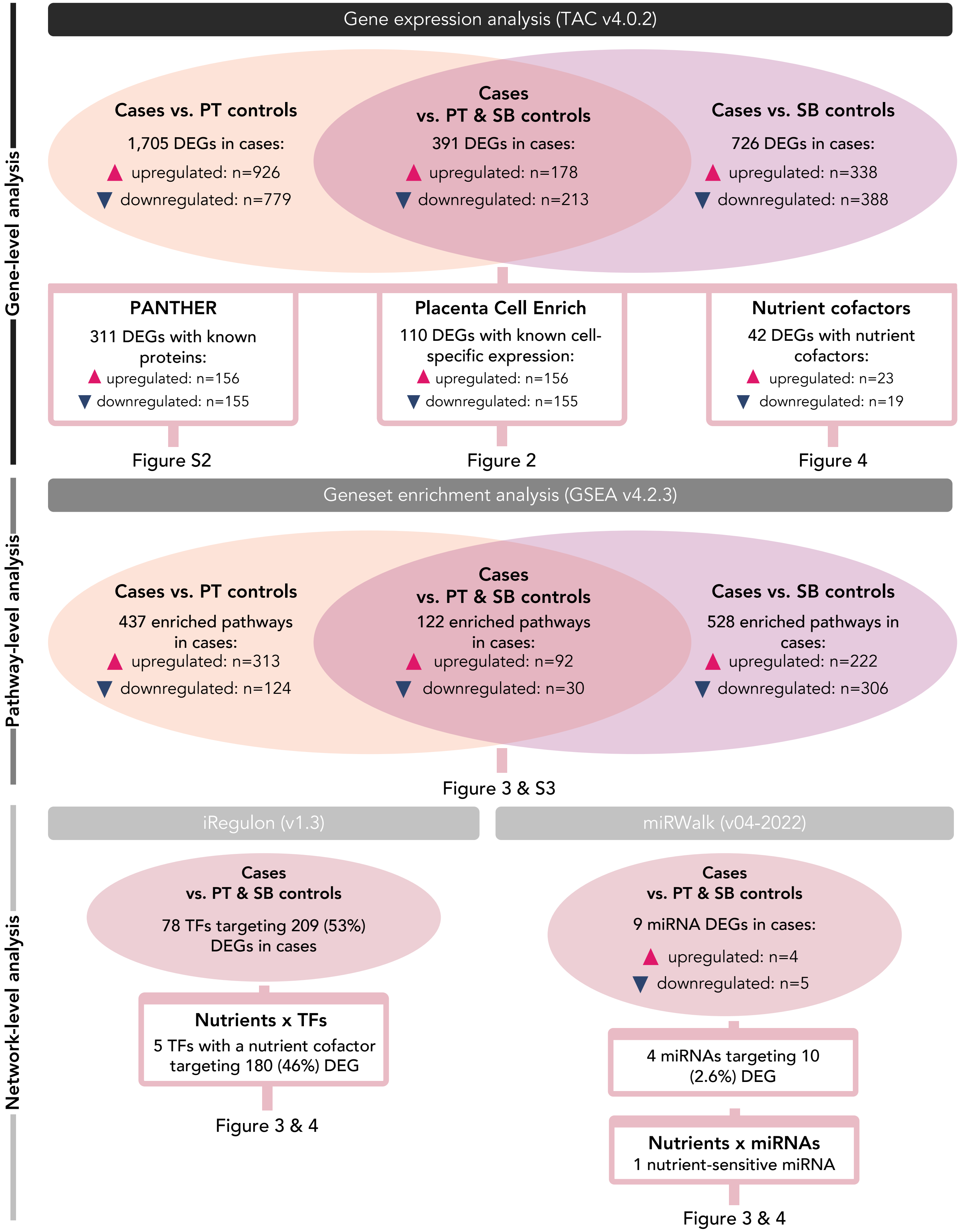


**Supplementary Figure S1. Transcriptome analysis workflow.** PT controls = preterm controls. SB controls = spina bifida study controls. DEG = differentially expressed (DE) genes (annotated; q<0.05 and FC≥2). TF = transcription factor.

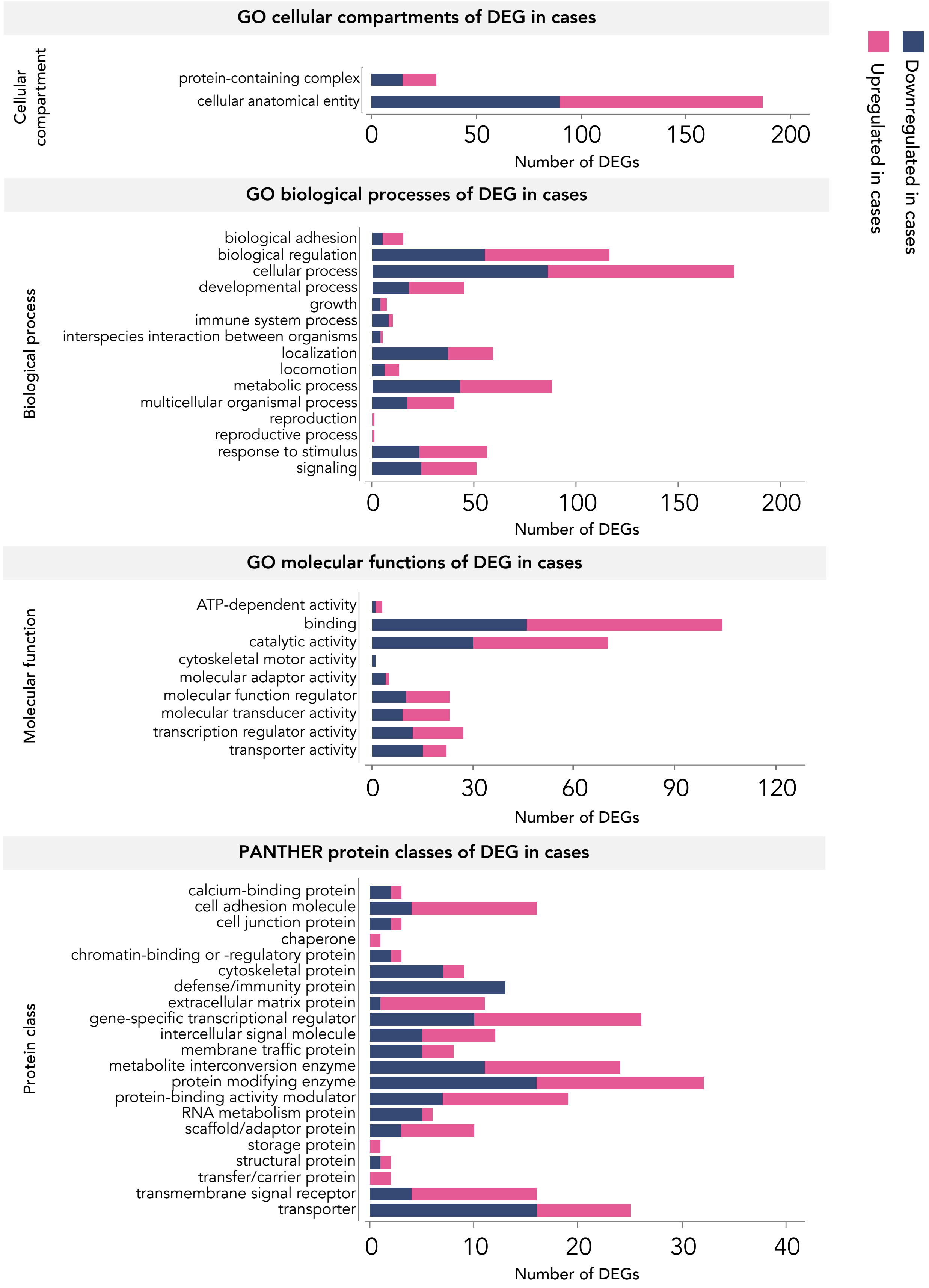


**Supplementary Figure S2. PANTHER annotations (cellular components, biological process, molecular function, and protein class) for differentially expressed genes in cases compared to PT and SB controls.** GO = gene ontology. DEG = differentially expressed genes. PANTHER = Protein ANalysis THrough Evolutionary Relationships.

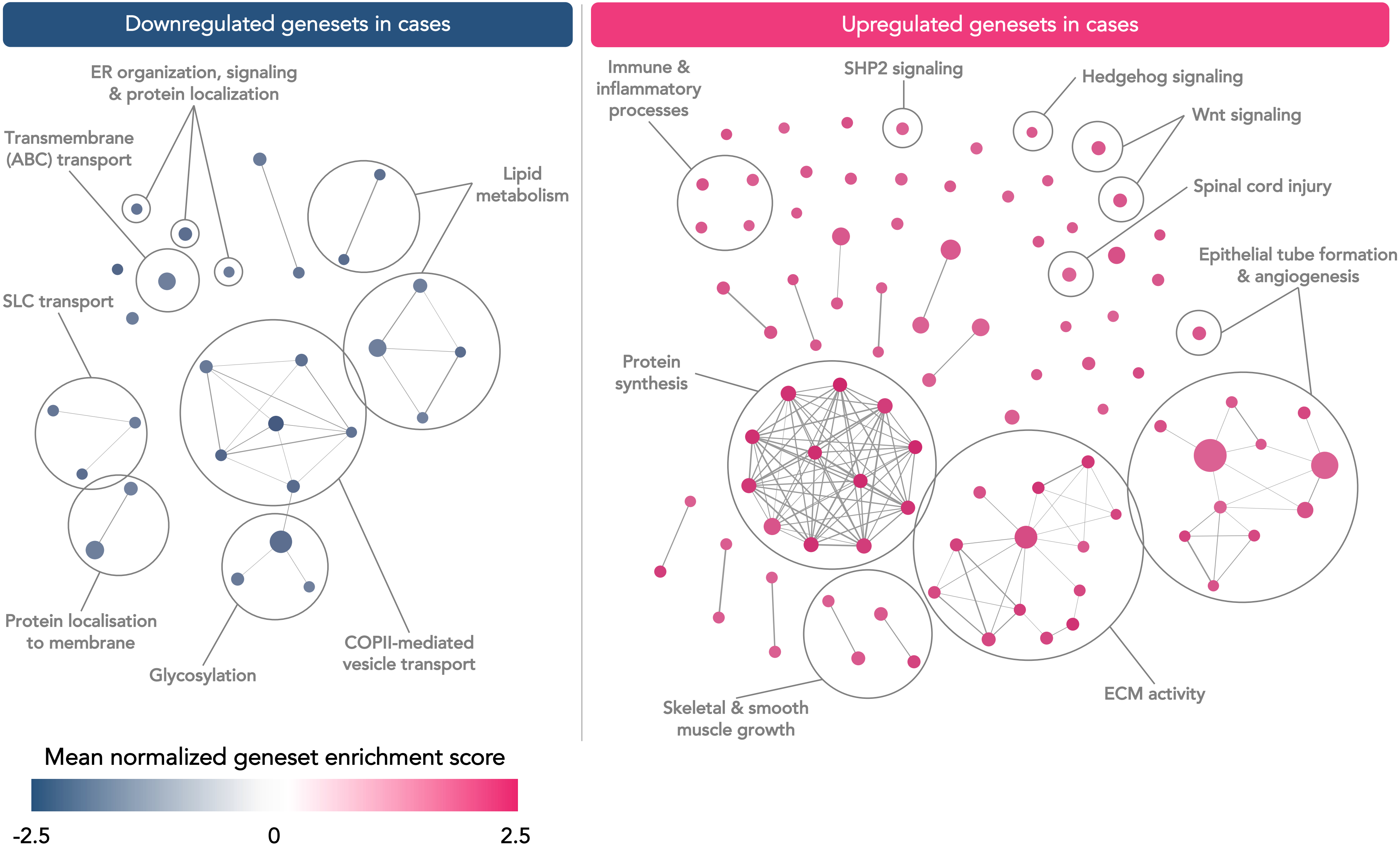


**Supplementary Figure S3. Enrichment map of differentially enriched genesets in case placentae (vs. PT and SB controls) with key annotations.** Geneset enrichment analysis revealed 122 enriched gene pathways (nodes) in cases compared to PT and SB controls (downregulated in cases: n=30 [blue nodes], upregulated in cases: n=92 [pink nodes]. Size of node = number of genes in pathway (17-530). Select genesets of interest are annotated here. Edge thickness = similarity coefficient (0-1). Solute carrier = SLC. ATP-binding cassette = ABC. Coat protein complex II = COPII. *SHP2* = SH2 containing protein tyrosine phosphatase-2. ECM = extracellular matrix.
