## Supplementary Tables S16-17 for "Fetal spina bifida associates with dysregulation in nutrient-sensitive placental gene networks: findings from a matched case-control study"

**Supplementary Table S16.** Maternal dietary recall data assessed as absolute intakes and dietary reference intakes for cases and SB controls.

| | SB controls (n=8) | | | Cases (n=10) | | | p value[a] |
|---|---|---|---|---|---|---|---|
| **Energy** | **Median (IQR)** | **% Below EER** | | **Median (IQR)** | **% Below EER** | | |
| Energy intake (kcal/day) | 1939 (1666, 2526) | 50 | | 1911 (1704, 2396) | 80 | | 0.94 |
| **Macronutrients** | | **% Below EAR** | **% Above TUL** | | **% Below EAR** | **% Above TUL** | |
| Carbohydrate (g/day) | 253 (167, 312) | 12.5 | - | 247 (216, 290) | 0 | - | 0.74 |
| Protein (g/day) | 83.8 (79.5, 102) | 0 | - | 69.6 (53.9, 89.6) | 20 | - | 0.19 |
| Total fat (g/day) | 76.7 (49.7, 106.3) | - | - | 76.2 (69.1, 92.4) | - | - | 0.77 |
| **Micronutrients** | | | | | | | |
| Calcium (mg/day) | 1080 (785, 1481) | 25 | 0 | 967 (691, 1121) | 30 | 0 | 0.23 |
| Choline (mg/day) | 340 (289, 553) | - | 0 | 190 (158, 381) | - | 0 | 0.045 |
| Copper (mg/day) | 1.35 (1.2, 2.39) | 0 | 0 | 1.25 (0.91, 1.6) | 10 | 0 | 0.12 |
| Iron (mg/day) | 13.3 (10.1, 18.9) | 100 | 0 | 12.3 (10.7, 16.7) | 90 | 0 | 0.89 |
| Magnesium (mg/day) | 313 (240, 458) | 37.5 | - | 293 (220, 420) | 60 | - | 0.66 |
| Niacin (mg/day) | 22.2 (16.8, 29.1) | 12.5 | - | 18.2 (13.7, 27.2) | 20 | - | 0.44 |
| Phosphorus (mg/day) | 1515 (1048, 1859) | 0 | 0 | 1253 (1047, 1749) | 0 | 0 | 0.4 |
| Potassium (mg/day) | 3811 (2339, 4194) | - | - | 2180 (1779, 2859) | - | - | 0.02 |
| Selenium (mcg/day) | 118 (78.3, 153) | 0 | 0 | 110 (78.7, 168) | 0 | 0 | 0.84 |
| Sodium (g/day) | 3.52 (2.62, 5.01) | - | 87.5 | 2.88 (2.07, 3.92) | - | 70 | 0.37 |
| Vitamin A (RAE, mcg/day) | 2158 (1094, 2501) | 0 | 0 | 865 (623, 1257) | 20 | 0 | 0.03 |
| Vitamin B1 (Thiamin; mg/day) | 1.6 (1.34, 1.78) | 0 | - | 1.51 (1.35, 1.77) | 10 | - | 0.82 |
| Vitamin B2 (Riboflavin; mg/day) | 2.09 (1.75, 3.1) | 0 | - | 1.91 (1.4, 2.16) | 20 | - | 0.14 |
| Vitamin B6 (mg/day) | 2.27 (1.31, 3.38) | 37.5 | 0 | 1.13 (1.06, 1.32) | 100 | 0 | 0.02 |
| Vitamin B9 (Folate; DFE, mcg/day)[b] | 436 (356, 645) | 75 | 0 | 465 (337, 566) | 60 | 0 | 1 |
| Vitamin B12 (mcg/day) | 6.25 (4.55, 7.42) | 12.5 | - | 2.69 (1.36, 4.48) | 50 | - | 0.007 |
| Vitamin C (mg/day) | 218 (124, 269) | 0 | 0 | 39 (24.4, 70.6) | 80 | 0 | 0.002 |
| Vitamin D (D2 + D3, mcg/day) | 7.59 (5.38, 11.3) | 75 | 0 | 2.61 (1.11, 4.63) | 100 | 0 | 0.004 |

| | | | | | | | |
|---|---|---|---|---|---|---|---|
| Vitamin E (mg/day) [c] | 13 (6.92, 17.1) | 37.5 | 0 | 7.93 (6.85, 10.7) | 80 | 0 | 0.12 |
| Zinc (mg/day) | 11.8 (10.1, 14.2) | 12.5 | 0 | 8.28 (6.98, 13.2) | 70 | 0 | 0.3 |
| **Overall nutrient EARs met (%)** | 81.6 (67.1, 84.2) | | | 55.3 (50, 67.1) | | | 0.01 |
| **Overall nutrient EARs met quartiles (n [%])** | | | | | | | 0.006 |
| Highest (80-95%) | 4 (50) | | | 0 | | | |
| Upper-middle (63-80%) | 3 (37.5) | | | 3 (30) | | | |
| Lower-middle (54-62%) | 0 | | | 5 (50) | | | |
| Lowest (32-53%) | 1 (12.5) | | | 2 (20) | | | |

[a] Data are median (IQR) with p value for one-way analysis of variance (ANOVA [normal distribution/equal variance]; Wilcoxon test [non-parametric data or outliers]) used to assess differences in average nutrient intake between groups. [b] The TUL consider folic acid intake only. [c] The TUL considers added vitamin E only. A "-" is used to indicate that no EAR or TUL was established for reference[1]. EER = estimated energy requirement. EAR = estimated average requirement. TUL = tolerable upper level. DFE = dietary folate equivalent. RAE = retinol activity equivalents.

**Supplementary Table S17.** Maternal Healthy Eating Index (HEI) and Dietary Inflammatory Index (DII) scores derived from ASA24 dietary recall.

| | Score range | SB controls (n=8) | Cases (n=10) | p value |
|---|---|---|---|---|
| | | Median (IQR) | Median (IQR) | |
| **HEI Dietary Components** | | | | |
| Adequacy score | 0-60 | 40.3 (29.1, 50.1) | | 0.06 |
| Total fruits | 0-5 | 5 (4.05, 5) | 1.08 (0.55, 2.34) | 0.02 |
| Whole fruits | 0-5 | 5 (4.25, 5) | 2.15 (1.05, 5) | 0.17 |
| Total vegetables | 0-5 | 4.9 (3.23, 5) | 2.23 (1.34, 3.38) | 0.047 |
| Greens and beans | 0-5 | 5 (1.11, 5) | 0.78 (0, 5) | 0.16 |
| Whole grains | 0-10 | 4.84 (0, 9.65) | 2.57 (0, 8.6) | 0.65 |
| Dairy | 0-10 | 9.3 (5.82, 10) | 6.88 (4.33, 9.55) | 0.26 |
| Total protein foods | 0-5 | 5 (4.47, 5) | 3.92 (2.74, 5) | 0.17 |
| Seafood and plant proteins | 0-5 | 1.15 (0, 5) | 3.78 (0, 5) | 0.78 |
| Fatty acids | 0-10 | 2.53 (0, 7.24) | 3.8 (0.87, 7.9) | 0.65 |
| Moderation score | 0-40 | 23.7 (17.9, 27.4) | 20.1 (15.2, 26.4) | 0.49 |
| Refined grains | 0-10 | 9.57 (7.03, 10) | 1.16 (0, 6.86) | 0.01 |
| Added sugars | 0-10 | 8.38 (6.12, 10) | 8.08 (6.31, 9.53) | 0.77 |
| Sodium | 0-10 | 4.29 (0, 4.82) | 5.01 (2.48, 8.53) | 0.14 |
| Saturated fats | 0-10 | 2.56 (0, 9.15) | 4.4 (3.2, 7.12) | 0.56 |
| Total score | 0-100 | 61.2 (48.3, 77) | 49.9 (43.6, 61) | 0.09 |
| HEI score rating (n [%]) | | | | 0.29 |
| Good | | 1 (11.1) | 0 | |
| Needs improvement | | 5 (55.5) | 5 (50) | |
| Poor | | 2 (22.2) | 5 (50) | |
| **Dietary inflammatory index** | | | | |
| Total DII score | | -0.95 (-2.3, 0.26) | 0.84 (0.44, 1.97) | 0.01 |
| DII score quartiles (n [%]) | | | | 0.04 |
| Highest (1.49-2.8) | | 1 (11.1) | 3 (30) | |
| Upper-middle (0.58-1.48) | | 1 (11.1) | 4 (40) | |
| Lower-middle (–0.9-0.57) | | 2 (22.2) | 3 (30) | |
| Lowest (–4.76- –0.89) | | 4 (44.4) | 0 | |

Data are median (IQR) with p value for one-way analysis of variance (ANOVA [normal distribution/equal variance]; Wilcoxon test [non-parametric data or outliers]) used to assess differences in HEI and DII scores. HEI = healthy eating index. DII = Dietary inflammatory index.
